# Sociodemographic and access-related correlates of sanitary pads among college students in Lucknow during Covid19

**DOI:** 10.1101/2020.10.14.20210815

**Authors:** Surbhi Garg, Ruqayya Alvi, Suman Gupta, Absar Ahmad

## Abstract

**Introduction:** Sanitary napkin is an essential aspect of the Menstrual management materials for women and adolescent girls between menarche and menopause. Despite being an important issue concerning women and girls in the menstruating age group, access to menstrual hygiene products neglected during the COVID19 pandemic. Further, there is no evidence of the practice of menstrual hygiene products in Indian settings during this period. This paper investigates the prevalence of socio-demographic correlations of access to sanitary napkins among college students in Lucknow.

**Methods:** An online retrospective cross-sectional survey was conducted in Lucknow in September 2020. In total, 1439 participants took part in the survey. After removing 55 participants, those quit the survey by clicking on the disagree button and 13 were not satisfying inclusion criteria. So the final samples were 1371, which were included in the analysis. Students of UG and PG currently studying in colleges in the Lucknow were eligible to participate. The data collection was anonymous. Responses were analysed using descriptive and bivariate logistic regression.

**Results:** In this study, 1371 students were included, making a response rate of 96.2 percent. Nearly 12.5 percent of students reported about difficulty encountered during the lockdown. Muslims, Father education illiterate or upto12th, father occupation as farmer, monthly salary less than 25 thousand, residence as rural, and history of reusable clothes were more likely to face problems to access sanitary pads during the lockdown in Lucknow (P < .05).

**Conclusions:** During COVID-19 lockdown, about 12.5 % of girls were dependent on either locally available resources as absorbents during menstruation or paid more to access in Lucknow. Because of the lockdown, many people have lost their livelihood. More than ever, economically low-income families are reluctant to spend on sanitary pads, which is why few college girls were going back to their previous handling periods by using rags.

**Highlights:**

- The prevalence of menstrual management products during Covid19 were unknown
- Women and Adolescents in India suffer the shortages of Sanitary napkin during lockdown
- Prevalence of access to sanitary pads should emphasis different focal points

## Introduction

It is a well-established fact that the COVID-19 and lockdown affected economically (1) and psychosocially (2) to all segments of the population adversely. Especially vulnerable groups, including people living below the poverty line, differently-abled people, women, children, elderly and migrant workers (3). Healthcare, a fundamental human right, also affected (4) because, it affected routine healthcare services due to shortages of essential medical supplies and health staff (5).

Across diverse social contexts, the topic of menstruation has often been relegated and it was not in priority list of policy makers(6).Global health priorities were aimed only at reducing maternal morbidity and mortality, and the problematic feminization of the HIV epidemic(7). Reason was because of menstrual shame and the complexities of menstrual management were perceived as an inevitable part of the social order, and other priorities for the limited existing resources consumed more attention(6).

However, now menstrual hygiene management (MHM) is one of globally recognized public health issue(6) which was also affected during COVID19 lockdown and made menstruating females one of vulnerable groups, because menstruation is a biological processes and do not cease during lockdown. With the sudden closure of campuses worldwide, many female students faced challenges to manage menstrual hygiene practices confined at home(8). The COVID-19 lockdown crisis was presented with even more difficulties in accessing menstrual hygiene products (9) in India, because it is not a socially discoursed area. There were lack of availability of sanitary pads (10), which facilitate personal hygiene, and dispose of its waste, which required adequate privacy(11). During lockdown around 58 per cent of the small and medium scale manufacturers of sanitary pads were not able to operate fully whereas 37 per cent were not operational at all(12).

Hygiene practices such as the use of sanitary napkins are useful during menstruation to protect girls or women health (13). The closure of shops and shutdown of transport means less availability and accessibility of menstrual hygiene provisions(14). Further, the lockdown was also challenging for women’s and girls’ access to toilets in presence of male and elder family members for managing menstruation (15).

India went into one of the world’s strictest lockdowns from March 25 (16) to May 31 (17), extended four times, from June 1 there were gradual relaxations. However, schools and colleges are still not open. During the initial phase of the lockdown, sanitary napkins were not included in the essential items, later rectified after massive public outrage. That led to a shortage and devised many girls and women with no choice but to resort to unhygienic practice using old clothes/rags to manage their periods (18).

Various State governments have a provision in a school-based distribution of sanitary pads under a central government programme, Kishori Shakti Yojna. Due to schools’ closing down, many girls have no access to sanitary pads forced to shift from disposable sanitary pads to cloth pads (18).

A common problem linked to cloth pads is that those who use it do not have adequate information on maintaining the cloth they are using so that they are not infected later. Using cloth pads without knowing how to keep it poses a danger to a woman’s health, and they risk infections of a grave nature (19).

Hygiene-related menstruation practices are essential as these can predispose women to the risk of reproductive tract infection (20). For menstruating girls and women, anxiety over menstrual hygiene is a key concern. The interplay of socio-economic status, menstrual hygiene practices, and RTI are noticeable (21). Women’s autonomy, residential status, occupation of father, Caste, exposure to mass media, and wealth status found to be the most significant positive factors of hygienic menstrual practices (20) (22).

National Family Health Survey (NFHS) 4 (2015 −16) of Uttar Pradesh reported, only 47.1% of women (Urban 68.6, Rural 39.9) in the age group of 15 to 24 years use hygienic methods of protection during their menstrual period (23). Girls with a higher education level use safe menstruation methods compare to less educated girls (24). So, college students better use of menstruation hygiene practices than the general women population. A study in Telangana reported about 87 % of college students using sanitary pads (25), while a study done in Ghana said 100% were using a sanitary pad (26).

Further, the lack of sanitary products created a sense of isolation and fear (27). In Lebanon, 69% of adolescent girls were walk long distance to access menstrual pads. Similarly, parents of 63% of young girl faced financial hardship to buy menstrual pads. In Uganda, 100% of respondents said they were struggling to access menstrual pads (28).

Menstrual hygiene is an important need of menstruating women and girls. Adolescent girls constitute a vulnerable group, particularly in India, where a girl child is neglected and discriminated(29). Menstruation is still regarded as something unclean or dirty among Indians (21). There is no study on menstrual management materials in India during the COVID-19 lockdown. Therefore, we conducted this study to know the prevalence of socio-demographic correlations of access to sanitary napkins among college students in Lucknow.

## Methods

### Study design and Settings

A quantitative approach was utilised to achieve the objectives of this study. An online cross-sectional survey was most appropriate for gathering information on access to sanitary napkins among college students between 11th September 2020 to 25th September 2020.

### Ethical approval

This work was done in collaboration of Shri Gurunanak Girls Degree College, Lucknow and Career Institute of Medical Sciences and Hospital, Lucknow. Ethical approval for this research was obtained from the Institutional Review Committee of Career Institute of Medical Sciences and Hospital (Ref: PHARMA/SEP/2020/02), Lucknow, India. Participants who gave consent to participate in the survey would click the ‘Agree’ button and then be directed to complete the self-administered questionnaire.

### Recruitment procedure

In total, 1439 participants took part in the survey. After removing participants’ data with 55 participants’, the questionnaires quit the survey by clicking on the disagree button and 13 not satisfying inclusion criteria. So the final samples were 1371, which were included in the analysis. As colleges were closed, it is not feasible to conduct a systematic sampling procedure; the researchers opted for Google form platform as an online survey. Students of UG and PG currently studying in colleges in the Lucknow were eligible to participate. We utilised several strategies to reach as many respondents as possible all over the Lucknow within the two-week data collection period. This includes relying on the researchers’ professional and personal networks to share the survey. WhatsApp was selected as the most popular communication and social platform during online teaching in colleges in Lucknow. Every college made a student’s group for communication-related to course instruction and notice. A description about the survey was given in the WhatsApp message postings before the link was provided to both English and Hindi language versions of the questionnaire.

### Measures

#### Outcome variables

Students were asked the following question: Did you face difficulties to access sanitary pads during lockdown? Response were recorded in No=0 and Yes=1. Further additional open question was also asked about alternative method used during that time.

#### Independent variable

We examined the distribution of this variable with a range of independent variables that have been suggested in the literature to predict menstrual absorbent precisely: Class of studying(*Graduation and Post-graduation*), Age(*<19,20-22,23+*) caste(*General, OBC, and SC/ST*), Religion(*Hindu, Muslim and Other*), Father education(*Illiterate or up to 12th, Graduate, and Postgraduate and higher*), Mother education(*Illiterate or up to 12th, Graduate and Postgraduate and higher*), father Occupation(*Farmer, Government employee, Self-employed/Businessman, Private employed, and Other*), Mother occupation(*Homemaker and working*), Monthly salary in Indian Rupees (*< 25 thousand, 25 thousand to 50 thousand, 50 thousand to 1 lakh and 1 lakh above*), Type of family(*Joint and Nuclear*), Place of residence(*Rural and Urban*) and history of reusable cloth use(*No and Yes*).

### Statistical analysis

For this study, the collected data were analysed using the Statistical Package for the Social Sciences (SPSS), version 25. To identify factors associated with difficulties to access sanitary pad during COVID19, we first performed bivariate analysis using Pearson’s chi-square test (χ^2^). All factors found significant at P-value<0.05 were incorporated into the bivariate logistic regression model. The statistical significance level was set at p < 0.05.

## Results

Table 1 depicts college students’ characteristics. The mean age of students was 20.06 years (SD = 1.78). There were more Undergraduate students (87.7%) participants, and the majority of students were Hindu (85.5%) followed by Muslims (11.7%). About 55.9% of students were General caste, followed by Other Backward Class (33.8%). About 48.8 percent of the student fathers were school educated, and 34.3 percent were graduate, followed by Postgraduate and higher (16.9%). Though 60.2 % of the student mothers were school educated, graduates (28%) and Postgraduate and higher (11.8%). The majority of student fathers were self-employed/businessman (27.9%) followed by private organization employees (26%), Government employees (20.7%), farmers (10%). Besides, 86.9% of student mothers were homemakers. The majority (59.1%) students’ monthly family’s income was less than 25 thousand and lived with a Nuclear family (66.5%). More than three fourth students’ place of residence was urban Lucknow. Nearly 15.1 % of students reported they had previously used reusable cloth.

**Table 1:** Descriptive statistics of college student participants in Lucknow(n=1371)

| **Variables** | **Categories** | **n** | **%** |
| --- | --- | --- | --- |
| Grade | *Graduation* | 1203 | 87.7 |
|  | *Post-Graduation* | 168 | 12.3 |
| Age | *<19* | 562 | 41.1 |
|  | *20-22* | 681 | 49.7 |
|  | *23 and above* | 128 | 9.3 |
| Caste | *General* | 767 | 55.9 |
|  | *OBC* | 463 | 33.8 |
|  | *SC/ST* | 141 | 10.3 |
| Religion | *Hindu* | 1172 | 85.5 |
|  | *Muslim* | 161 | 11.7 |
|  | *Other* | 38 | 2.8 |
| Father education | *School education* | 669 | 48.8 |
|  | *Graduate* | 470 | 34.3 |
|  | *PG and above* | 232 | 16.9 |
| Mother education | *School education* | 825 | 60.2 |
|  | *Graduate* | 384 | 28.0 |
|  | *PG and above* | 162 | 11.8 |
| Father occupation | *Farmer* | 137 | 10.0 |
|  | *Government employee* | 284 | 20.7 |
|  | *Self-Employed/Businessman* | 383 | 27.9 |
|  | *Private Org. employee* | 357 | 26.0 |
|  | *Other* | 210 | 15.3 |
| Mother occupation | *Housewife* | 1191 | 86.9 |
|  | *Working* | 180 | 13.1 |
| Monthly salary(INR) | *<25 thousand* | 809 | 59.1 |
|  | *25 to 50 thousands* | 311 | 22.7 |
|  | *50 thousands to 1 lakh* | 163 | 11.9 |
|  | *1 lakh and above* | 88 | 6.4 |
| Type of family | *Joint* | 459 | 33.5 |
|  | *Nuclear* | 912 | 66.5 |
| Place of Residence | *Rural* | 319 | 23.3 |
|  | *Urban* | 1052 | 76.7 |
| History of uses of reusable cloth | *No* | 1164 | 84.9 |
|  | *Yes* | 207 | 15.1 |
| **Total** |  | **1371** | **100** |

Source: Online primary survey

Table 2 exhibits the relationship between students’ characteristics and difficulties to access sanitary pad during COVID19 lockdown. The present study reported nearly 12.5 percent of students encountered a problem to access sanitary pad during the lockdown. Caste, religion, Father’s education, Mother’s education, Father’s occupation, Mother’s occupation, Monthly salary, type of family, residence place, and history of reusable cloth uses were statistically associated with difficulties to access sanitary pads during COVID19 lockdown.

**Table 2:**
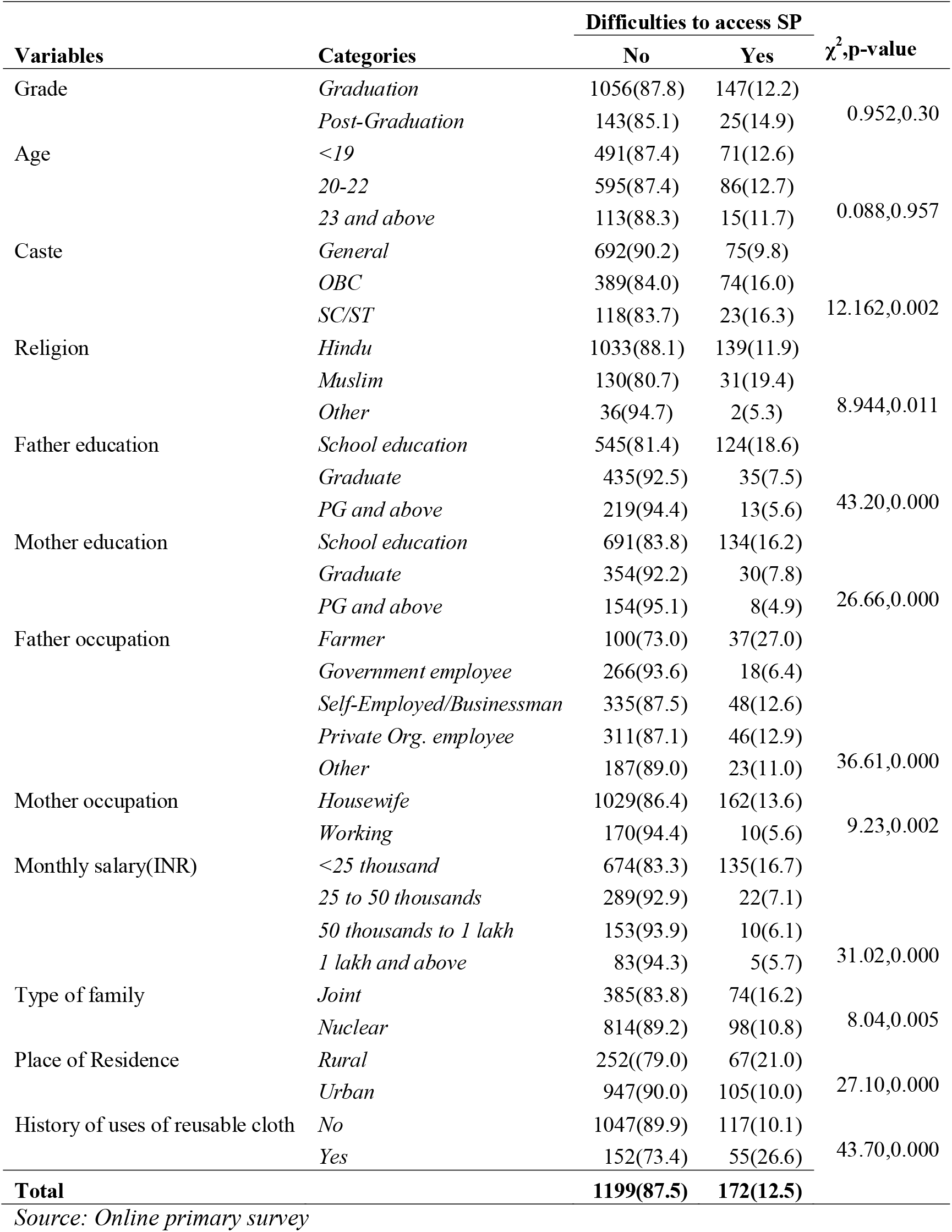
Study variables and bivariate analysis examining factors associated with difficulties to access sanitary pad(SP) during Covid19 lockdown among college students in Lucknow (*n=1371*)

Difficulties encountered to access sanitary pad access during the COVID19 lockdown were influenced significantly by religion, father’s education, father’s occupation, monthly salary, place of residence, and history of reusable cloth use. Among religion, Muslim students were more likely (AOR 2.182 95%CI:1.354-3.517) to encounter difficulties in accessing sanitary pads compared to Hindu students. Furthermore, Students whose fathers were graduates were less likely (AOR 0.559 95% CI:0.349-0.897) to encounter difficulties accessing sanitary pads than those whose fathers were school educated. Students whose father were farmers were more likely (AOR 1.998 95% CI:1.013-3.937) face difficulties accessing sanitary pads than those whose fathers were government employees. Students whose monthly family income was between 25 to 50 thousand were less likely (AOR 0.560 95% CI:0.334-0.937) encounter difficulties to access sanitary pads compare to those whose monthly income was less than 25 thousand. Students’ residence location was rural and more likely (AOR 1.736 95%CI: 1.194-2.525) meet difficulties accessing sanitary pads compared to their counterparts. Students with a history of reusable clothes were more likely to face problems (AOR 2.858 CI:1.942-4.205) to access sanitary pads compared to those who have not previously used reusable cloth (Table 3).

**Table 3:** Binary logistic regression analysis of predictors of difficulties to access sanitary pads during covid19 lockdown by background characteristics of college students in Lucknow

| Variables | Categories | Unadjusted OR<br>(95%C.I.) | Adjusted OR<br>(95% C.I.) |
| --- | --- | --- | --- |
| Caste | *General*<sup>®</sup> |  |  |
|  | *OBC* | 1.755(1.244-2.477)** | 1.208(0.827-1.765) |
|  | *SC/ST* | 1.798(1.084-2.984)* | 1.215(0.687-2.147) |
| Religion | *Hindu*<sup>®</sup> |  |  |
|  | *Muslim* | 1.772(1.153-2.724)** | 2.182(1.354-3.517)** |
|  | *Other* | 0.413(0.098-1.734) | 0.542(0.122-2.42) |
| Father education | *School education*<sup>®</sup> |  |  |
|  | *Graduate* | 0.354(0.238-0.525)** | 0.559(0.349-0.897)* |
|  | *PG and above* | 0.261(0.144-0.472)** | 0.498(0.241-1.032) |
| Mother education | *School education*<sup>®</sup> |  |  |
|  | *Graduate* | 0.437(0.288-0.663)** | 0.844(0.505-1.41) |
|  | *PG and above* | 0.268(0.129-0.558)** | 0.873(0.357-2.136) |
| Father occupation | *Government employee*<sup>®</sup> |  |  |
|  | *Farmer* | 5.468(2.976-10.047)** | 1.998(1.013-3.937)* |
|  | *Self-Employed/Businessman* | 2.117(1.203-3.726)** | 1.289(0.701-2.373) |
|  | *Private Org. employee* | 2.186(1.237-3.861)** | 1.45(0.78-2.695) |
|  | *Other* | 1.818(0.954-3.463) | 0.987(0.488-2.00) |
| Mother occupation | *Housewife*<sup>®</sup> |  |  |
|  | *Working* | 0.374(0.193-0.722)** | 0.714(0.348-1.464) |
| Monthly salary(INR) | *<25 thousand*<sup>®</sup> |  |  |
|  | *25 to 50 thousands* | 0.380(0.237-0.609)** | 0.56(0.334-0.937)* |
|  | *50 thousands to 1 lakh* | 0.326(0.168-0.635)** | 0.541(0.265-1.107) |
|  | *1 lakh and above* | 0.301(0.120-0.756)* | 0.489(0.184-1.302) |
| Type of family | *Joint*<sup>®</sup> |  |  |
|  | *Nuclear* | 0.626(0.452-0.867)** | 0.761(0.534-1.083) |
| Place of Residence | *Urban*<sup>®</sup> |  |  |
|  | *Rural* | 2.398(1.713-3.357)** | 1.736(1.194-2.525)** |
| History of uses of reusable cloth | *No*<sup>®</sup> |  |  |
|  | *Yes* | 3.238(2.253-4.654)** | 2.858(1.942-4.205)** |

®Reference category, OR: Odds Ratio, \*Significant at p<0.05, \*\*Significant at p<0.01
Source: Online primary survey

## Discussion

This is the first study to evaluate difficulties to access sanitary pads during COVID19 lockdown in any countries. Our study is only focused on college students especially Undergraduate and Postgraduate students in Lucknow which is capital of Uttar Pradesh state in India. Data also shows that Lucknow is the worst COVID-19 affected district in UP, a fifth-worst COVID-19 affected state in India (30). Since the COVID-19 outbreak media reported about difficulties for menstruating women in accessing menstrual hygiene products (31) in India as well as abroad(32)(33). However, there has been no scientific evidence available of the difficulties faced by the menstruating women to access sanitary pads during lockdown. This study is also small steps to add a new evidence about the prevalence of difficulties as well as predictors to access sanitary pads.

Present study found 12.5 percent college students faced difficulties to access sanitary pads during lockdown. The commonly used alternative for pads during lockdown were homemade reusable cloth pad. Similar type of pad were also used in Rwanda, however, women and girls also used rags, leaves, newspapers or other makeshift items to absorb or collect menstrual blood(34). Most girls in India is not comfortable for asking men in the family to procure pads for them since periods is a taboo subject not openly talked about in Indian families. In addition, during lockdown when all the men were at home because of the lockdown, girls and women may find it difficult to use toilet facilities as frequently and getting additional water to wash pads and put them out in the sun to dry (10). Students managed menstruation during lockdown by using the pad of brand which they never used earlier and size, or by paid more price, bought from another place, used old cloth, home-made pad, and sponge.

Present study revealed that Muslims students faced difficulties to access sanitary pads during COVID19 lockdown. Reason may be Muslims in India displayed lowest work participation rate(35). Further, Muslims mostly rely on the informal economy (36) and during COVID19 lockdown informal workers suffer more (37) in India as well as abroad(38). It may be the reason because of low family income and poor income during lockdown, Muslim girls may compromise with their menstrual needs and personal hygiene.

Students whose fathers were graduates were less likely to encounter difficulties accessing sanitary pads than those whose fathers were school educated. In previous study in Nepal also reported association of good menstrual hygiene practice with father’s education(39).Educated father were have more awareness and concerned about the hygiene of their daughter.

Students whose father were farmers as well rural residence students were more likely face difficulties accessing sanitary pads compared to those whose father were government employees. In previous studies in India(20) and Ghana(40) where adolescents whose fathers were farmers had poor menstrual hygiene management were found. During COVID-19 lockdown time Indian farmers faced new challenges in front of agriculture sector that is already under threat. The lockdown created both a shortage of labour, equipment, access to seeds, fertilizers and pesticides (41).This all make farmers highly vulnerable to the present crisis. Reason In rural areas, women do not have access to sanitary products so uses of sanitary napkins is very low. Another reasons were high cost of branded napkins, and cultural barriers (42). So, they mostly rely on reusable cloth pads which they wash and use again(43). However, college students purchase it when they come to college. But due to lockdown they are not able to come outside.

Students whose monthly family income was between 25 to 50 thousand were less likely to encounter difficulties accessing sanitary pads than those whose monthly income was less than 25 thousand. Due to limited household income during the lockdown, people prioritized other household demands and did not prefer to spend their fixed income purchasing sanitary pads. On the other hand, prosperous people buy the product in mass. In India society is patriarchal and during lockdown women are not able to told their family member for sanitary napkin.

Men do not support women regarding menstruation hygiene and do not give money to buy menstrual products. So women have to rely on cheap reusable cloth pads which they have to wash, dry, and use again.

Study also found students with a history of reusable clothes were more likely to face problems to access sanitary pads compared to those who have not previously used reusable cloth. They may more chance that students those unable to afford such products due to high cost previously may also not afford it during lockdown any may be from lower income or rural areas. So, they mostly rely on reusable cloth pads which they wash and use again(43).

### Strength and limitations of the study

The study tried to evaluate the difficulties to access sanitary pads during lockdown by online survey and tried to cover all types of students. However, this study has a limitation. First, the study design’s cross-sectional nature might not show the cause and effect relationships between study variables. Second, this study follows only quantitative data collection, and mixed approaches do not triangulate it. Third, the study lacks random sampling, which leads the statistical confidence and margin of error.

## Conclusion

About 12.5 college students faced difficulties with accessing sanitary pads during the lockdown. Muslims, Father education illiterate or upto12th, Father occupation as farmer, Monthly salary less than 25 thousand, place of residence as rural, and history of uses of reusable clothes were more likely to face problems to access sanitary pads were significant correlates of difficulties to access sanitary pads during the lockdown in Lucknow. It demonstrates a need to design acceptable advocacy programs for menstruating females during a crisis because menstruation does not cease during any emergency.

## Data Availability

This research is based on primary data.

## Acknowledgment

We thank all the faculties of various colleges of Lucknow to motivate their students to participate in the studies

## Notes

### Competing Interest Statement

The authors have declared no competing interest.

### Funding Statement

Not received any funding

### Author Declarations

This work was done in collaboration of Shri Gurunanak Girls Degree College, Lucknow and Career Institute of Medical Sciences and Hospital, Lucknow. Ethical approval for this research was obtained from the Institutional Review Committee of Career Institute of Medical Sciences and Hospital, Lucknow, India.

